# Urogenital Schistosomiasis Among Pre-School and School-Age Children During a Community-Based Control Programme in the Lake Chad Basin: Implications for Preventive Chemotherapy and Integrated Control

**DOI:** 10.64898/2026.07.28.26359151

**Authors:** Didier Lalaye, Marius Madjissem, Loukoum Nan-Arabé

## Abstract

**Background:** Urogenital schistosomiasis control programmes have traditionally focused preventive chemotherapy (PC) on school-aged children (SAC, 5–14 years). However, the burden of infection among pre-school-aged children (Pre-SAC) and the extent to which infection patterns differ from those of adults remain poorly documented in the Lake Chad Basin. This study presents a secondary analysis of routinely collected programme data from the Dawa Mobile Health schistosomiasis control programme to characterise the burden, age distribution, and determinants of urogenital schistosomiasis among children and adolescents (1–20 years) in the Ngouri Health District, Chad.

**Methods:** We conducted a retrospective cross-sectional secondary analysis of routinely collected data collected during a community-based schistosomiasis control programme implemented in the Ngouri Health District, Lac Province, Chad, between February 2024 and April 2025. All individuals screened during routine programme activities were eligible for inclusion. Children and adolescents aged 1–20 years (n = 1,946) constituted the primary population of interest, while adults aged ≥21 years (n = 2,570) served as a comparison group. Urogenital schistosomiasis was diagnosed by urine membrane filtration, with infection defined as the detection of at least one *Schistosoma haematobium* egg per 10 mL of urine. Multivariable logistic regression was used to identify factors independently associated with infection among children and adolescents.

**Results:** Among 4,516 programme participants, children and adolescents represented 43.1% of those screened and accounted for 43.9% of the 880 confirmed infections. During programme implementation, the observed prevalence was 19.8% (95% CI: 18.1–21.7%) among children and adolescents, 18.2% (95% CI: 14.7–22.2%) among Pre-SAC (1–10 years), and 20.3% (95% CI: 18.3–22.4%) among SAC (11–20 years), with no significant difference compared with adults (19.2%; *p* = 0.633). Both child age groups remained within the WHO moderate-endemicity category, indicating continued eligibility for annual preventive chemotherapy. In multivariable analysis, neither age subgroup, sex, education level nor occupation was independently associated with infection (all *p* > 0.05), suggesting broadly distributed community exposure.

**Conclusions:** Pre-school-aged children experienced a burden of urogenital schistosomiasis comparable to that of school-aged children and adults in this agro-pastoral setting. Although the reported prevalence reflects infection observed during programme implementation rather than baseline district prevalence, moderate endemicity persisted after 15 months of community-based screening and treatment, supporting the continuation of community-wide preventive chemotherapy, including Pre-SAC. These findings also suggest that chemotherapy alone may be insufficient to interrupt transmission and highlight the need to complement treatment with strengthened WASH interventions and adaptive surveillance strategies capable of identifying persistent transmission hotspots.

## INTRODUCTION

Urogenital schistosomiasis caused by *Schistosoma haematobium* remains one of the most widespread neglected tropical diseases in sub-Saharan Africa, affecting an estimated 112 million people, most of whom are children. The World Health Organization (WHO) recommends preventive chemotherapy (PC) with praziquantel as the cornerstone of morbidity control, with treatment strategies guided primarily by the prevalence observed among school-aged children (SAC). According to WHO guidelines, communities where SAC prevalence ranges from 10% to <50% require annual mass drug administration (MDA), whereas prevalence ≥50% warrants biannual treatment [1,2].

Although this strategy has substantially reduced schistosomiasis-related morbidity, growing evidence indicates that limiting surveillance and treatment to school-aged children may overlook other important reservoirs of infection. In particular, pre-school-aged children (Pre-SAC) are increasingly recognised as being exposed to infested water from an early age and may harbour infection burdens comparable to those of older children in many endemic settings [3,4]. In the agro-pastoral communities of the Lake Chad Basin, where daily activities such as water collection, livestock watering, fishing, and agriculture involve entire households, exposure to transmission sites begins well before school enrolment. Consequently, excluding younger children from surveillance and treatment programmes may reduce the overall effectiveness of schistosomiasis control.

Another important but insufficiently documented question concerns the age distribution of infection within endemic communities. If schistosomiasis affects children and adults similarly, restricting preventive chemotherapy to school-aged children alone may leave a substantial proportion of infected individuals untreated, thereby sustaining transmission. Conversely, if infection is concentrated in specific age groups, targeted interventions may remain appropriate [5]. Understanding how infection is distributed across the life course is therefore essential for optimising control strategies and interpreting WHO treatment recommendations.

The Ngouri Health District, located in Chad’s Lac Province, has historically been recognised as an endemic area for urogenital schistosomiasis, with a baseline prevalence of approximately 33.1% before implementation of community control activities [6]. Since February 2024, the Dawa Mobile Health programme has implemented routine community-based screening and treatment activities across villages in the district. During the 15-month implementation period, epidemiological and clinical data were systematically collected as part of routine programme activities rather than through a protocol-driven epidemiological study [7]. These routinely collected data provide an opportunity to assess the burden and age distribution of infection observed during programme implementation while recognising that the estimated prevalence does not represent the baseline prevalence of the district.

Importantly, despite 15 months of community-based screening and treatment, transmission may remain sufficiently high to warrant continued annual preventive chemotherapy under current WHO recommendations. If so, this would suggest that preventive chemotherapy alone is unlikely to interrupt transmission in highly endemic agro-pastoral settings and that complementary interventions—including improved water, sanitation and hygiene (WASH), strengthened health education, and adaptive surveillance approaches such as dynamic spatial risk mapping—may be required to optimise long-term control.

The present study therefore reports a secondary analysis of routinely collected programme data to: (1) estimate the observed period prevalence and disease burden of urogenital schistosomiasis among Pre-SAC (1–10 years) and SAC (11–20 years); (2) compare infection patterns between children and adults (≥21 years); (3) evaluate the observed prevalence against WHO endemicity thresholds for preventive chemotherapy; and (4) identify sociodemographic factors associated with infection among children and adolescents. Rather than estimating the underlying prevalence of the Ngouri Health District, this study describes infection patterns observed during programme implementation and discusses their implications for future schistosomiasis control strategies.

## MATERIALS AND METHODS

### Study Design and Setting

This study is a retrospective cross-sectional secondary analysis of routinely collected programme data generated through the Dawa Mobile Health schistosomiasis control programme implemented in the Ngouri Health District, Lac Province, Chad, between February 2024 and April 2025. The programme comprised community-based screening, parasitological diagnosis, and treatment of urogenital schistosomiasis delivered as part of routine public health activities. The research question and statistical analysis plan were developed after completion of programme implementation using the existing programme database.

The primary population of interest consisted of children and adolescents aged 1–20 years. Adults aged 21 years and above were included as a comparison group to examine age-related patterns of infection. A detailed description of the study area, district characteristics, and operational organisation of the Dawa Mobile Health programme has been reported elsewhere [6].

### Age Group Definitions

Age categories were defined according to the grouped age information available in the programme database and aligned, where possible, with WHO programmatic classifications for schistosomiasis control [2]:

- **Pre-school and early school-age children (Pre-SAC):** 1–10 years
- **School-age children and adolescents (SAC):** 11–20 years
- **Young adults:** 21–30 years
- **Adults:** 31–40 years
- **Older adults:** ≥41 years

Throughout this paper, the term *children and adolescents* refers to participants aged 1–20 years. Because age was recorded in grouped categories during routine programme implementation, the 1–10-year category includes both pre-school children (1–4 years) and early school-age children (5–10 years), preventing further age-specific disaggregation.

### Participant Enrolment and Data Collection

The Dawa Mobile Health programme provided voluntary community-based screening in participating villages throughout the implementation period. All individuals presenting for screening were eligible for inclusion. Children were enrolled following oral informed consent provided by a parent or legal guardian.

As part of routine programme activities, demographic and clinical information was recorded, including age group, sex, education level, and occupation. A urine specimen was collected from every participant for parasitological examination. Because this study was based on routinely collected programme data, participant enrolment and data collection were conducted for programme implementation rather than for the purposes of the present analysis.

### Diagnostic Procedures

Urine specimens were collected between 09:00 and 14:00 hours, corresponding to the period of peak *Schistosoma haematobium* egg excretion [8]. Body weight was measured in children to determine weight-based praziquantel dosing.

Samples were analysed using the WHO-recommended membrane filtration technique (polycarbonate membrane, 12–20 μm pore size), followed by microscopic examination for *S. haematobium* eggs [9]. Participants were considered infected when at least one *S. haematobium* egg was detected per 10 mL of urine.

As part of routine data quality assurance, a biological consistency check was performed. Twenty-two participants presenting with visible haematuria but initially recorded as parasitologically negative were reviewed and recoded as positive because of the high biological specificity of macroscopic haematuria for urogenital schistosomiasis in this epidemiological setting. All confirmed positive participants received praziquantel (40 mg/kg body weight) free of charge in accordance with national programme procedures.

### WHO Endemicity Classification

Observed prevalence estimates were interpreted according to WHO endemicity thresholds for preventive chemotherapy [2]: low endemicity (<10%), moderate endemicity (10% to <50%), and high endemicity (≥50%). These thresholds were used to assess the programmatic implications of the observed prevalence within each age group.

Because the data were collected during an ongoing community-based control programme, the reported prevalence should be interpreted as prevalence observed during programme implementation rather than as the baseline prevalence of the Ngouri Health District.

### Statistical Analysis

Observed period prevalence was calculated for each age group with 95% confidence intervals using the Wilson score method. Differences in prevalence between children and adults, and between Pre-SAC and SAC, were assessed using Pearson’s chi-square test. Crude odds ratios (ORs) were estimated using univariable logistic regression. The contribution of each age group to the overall burden of disease was expressed as the proportion of all confirmed cases.

A multivariable logistic regression model was fitted among children and adolescents (1–20 years) to identify factors independently associated with parasitological positivity. Pre-specified covariates included age subgroup (Pre-SAC versus SAC), sex, and education level. Model performance was assessed using McFadden’s pseudo-*R*².

All statistical analyses were performed using Python version 3.11 (statsmodels version 0.14 and SciPy version 1.11). Statistical significance was defined as a two-sided *p* value <0.05.

### Ethical Considerations

The Dawa Mobile Health programme was approved by the Scientific Ethics Committee of Chad and the National Bioethics Committee of Chad before programme implementation. Oral informed consent was obtained from all adult participants and from the parents or legal guardians of participating children in accordance with routine programme procedures.

The present study consisted of a secondary analysis of anonymised programme data. No additional participant contact or data collection was undertaken specifically for this analysis.

## RESULTS

### Study Population and Age Distribution

A total of 4,516 individuals screened through the Dawa Mobile Health programme between February 2024 and April 2025 were included in this secondary analysis. Children and adolescents aged 1–20 years accounted for 1,946 participants (43.1%), while adults (≥21 years) represented 2,570 participants (56.9%). During the programme period, the overall observed prevalence of urogenital schistosomiasis was 19.5% (880/4,516; 95% CI: 18.4–20.7%) after biological consistency correction. The study population distribution and absolute case burden by age group are presented in Table 1 and Figure 1.

**Figure 1.**
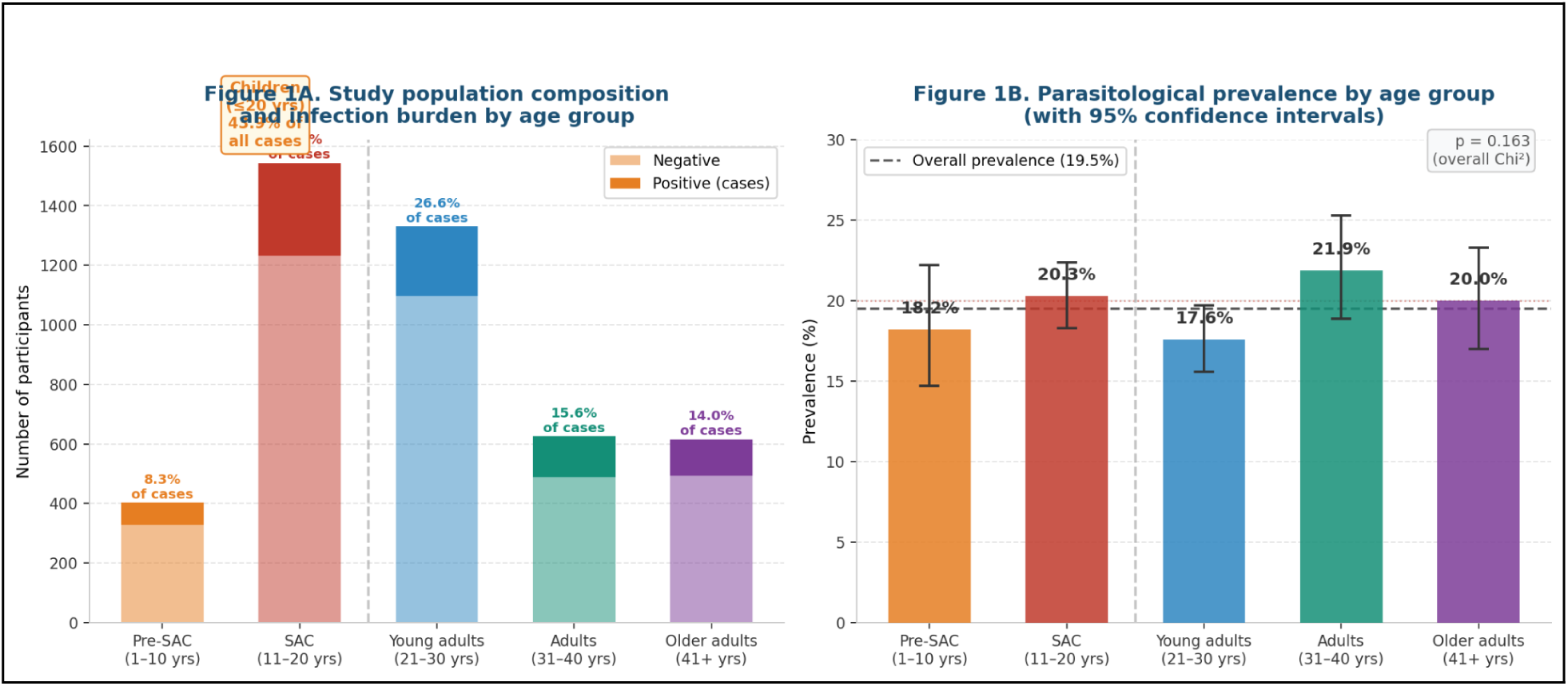
Disease burden and parasitological prevalence of urogenital schistosomiasis by age group (N = 4,516) Left (1A): Stacked bar chart showing total enrolled participants and positive cases by age group. Children and adolescents (1–20 years) account for 43.9% of all confirmed cases. Right (1B): Parasitological prevalence (%) with 95% confidence intervals by age group. Dashed line = overall period prevalence (19.5%). The vertical dashed line separates children/adolescents from adults. p = 0.163 (overall Chi², df = 4).

**Figure 2.**
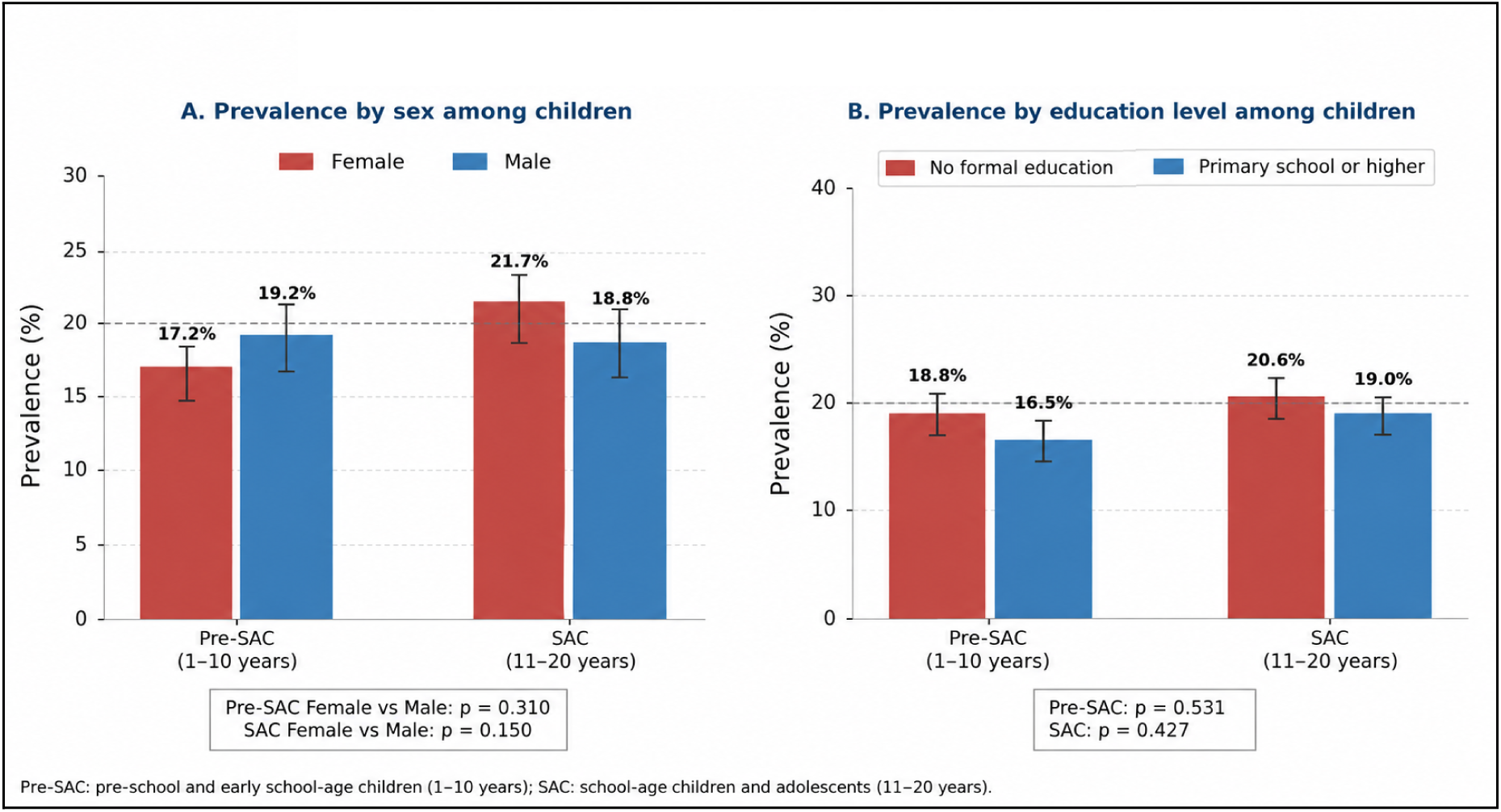
Prevalence of urogenital schistosomiasis among children by sex and education level. **Panel A:** Observed prevalence of urogenital schistosomiasis by sex among Pre-SAC (1–10 years) and SAC (11–20 years), with 95% confidence intervals. No statistically significant difference in prevalence was observed between males and females in either age group. **Panel B:** Observed prevalence according to education level among children and adolescents (1–20 years). Prevalence was similar across education categories, with no evidence of an association between educational attainment and infection.

**Table 1.** Study population, parasitological prevalence, and disease burden by age group (N = 4,516)

| Age group | N | % of total | Positive cases | Prevalence (%) | 95% CI | % of all cases |
| --- | --- | --- | --- | --- | --- | --- |
| Pre-SAC (1–10 years) | 402 | 8.9 | 73 | 18.2 | 14.7–22.2 | 8.3 |
| SAC (11–20 years) | 1,544 | 34.2 | 313 | 20.3 | 18.3–22.4 | 35.6 |
| **Children & adolescents (1–20 years)** | **1,946** | **43.1** | **386** | **19.8** | **18.1–21.7** | **43.9** |
| Young adults (21–30 years) | 1,330 | 29.5 | 234 | 17.6 | 15.6–19.7 | 26.6 |
| Adults (31–40 years) | 625 | 13.8 | 137 | 21.9 | 18.9–25.3 | 15.6 |
| Older adults (41+ years) | 615 | 13.6 | 123 | 20.0 | 17.0–23.3 | 14.0 |
| **All adults (21+ years)** | **2,570** | **56.9** | **494** | **19.2** | **17.8–20.8** | **56.1** |
| **Total** | **4,516** | **100.0** | **880** | **19.5** | **18.4–20.7** | **100.0** |

Pre-SAC: pre-school and early school-age children (1–10 years); SAC: school-age children and adolescents (11–20 years). Prevalences estimated by Wilson score method. Cases corrected for biological consistency (22 haematic-negative recoded as positive).

### Prevalence in Children vs. Adults

The period prevalence among all children and adolescents (19.8%; 95% CI: 18.1%–21.7%) was virtually identical to that observed among adults (19.2%; 95% CI: 17.8%–20.8%), with no statistically significant difference (Chi² = 0.23; df = 1; **p = 0.633**; OR = 1.04; 95% CI: 0.88–1.22). This pattern was consistent between the two child subgroups: Pre-SAC prevalence (18.2%; 95% CI: 14.7%–22.2%) was not significantly different from SAC prevalence (20.3%; 95% CI: 18.3%–22.4%; p = 0.357) or from adult prevalence (p = 0.637).

### WHO Endemicity Classification

Although these estimates were obtained during programme implementation rather than from a baseline epidemiological survey, both Pre-SAC and SAC remained within the WHO moderate-endemicity category, indicating that annual preventive chemotherapy would still be recommended. Neither group met the threshold for high endemicity (≥50%) that would justify biannual treatment. Importantly, the Pre-SAC prevalence (18.2%) substantially exceeds the low-endemicity threshold of 10%, establishing that this group requires inclusion in PC programs. These classifications are compared against WHO thresholds in Figure 4.

### Sex Distribution Among Children

Among children and adolescents, females represented 50.9% of the enrolled population (n=991) and males 49.1% (n=955). Prevalence was 20.8% (95% CI: 18.4%–23.4%) in females and 18.8% (95% CI: 16.5%–21.5%) in males, with no statistically significant difference (p = 0.310). In the Pre-SAC subgroup, prevalence was 17.2% in girls vs. 19.2% in boys (p > 0.05). Among SAC, prevalence was 21.7% in girls vs. 18.8% in boys (p = 0.150). No sex-based differential was observed in any child subgroup.

### Education Among Children

Among Pre-SAC children, 64.6% had no formal schooling (n=260), while 27.1% were attending primary school (n=109). Among SAC, 64.7% reported no schooling (n=999).

### Multivariable Logistic Regression — Children and Adolescents

A pre-specified multivariable logistic regression model was fitted among children and adolescents (n=1,911 after exclusion of non-standard entries). After simultaneous adjustment for age subgroup, sex, and education level, no variable reached statistical significance (Table 3, Figure 3). The McFadden pseudo-R² of 0.003 confirms that the pre-specified variables collectively explain a negligible proportion of variance in infection status among children, consistent with the homogeneous distribution of infection observed across all subgroups.

**Figure 3.**
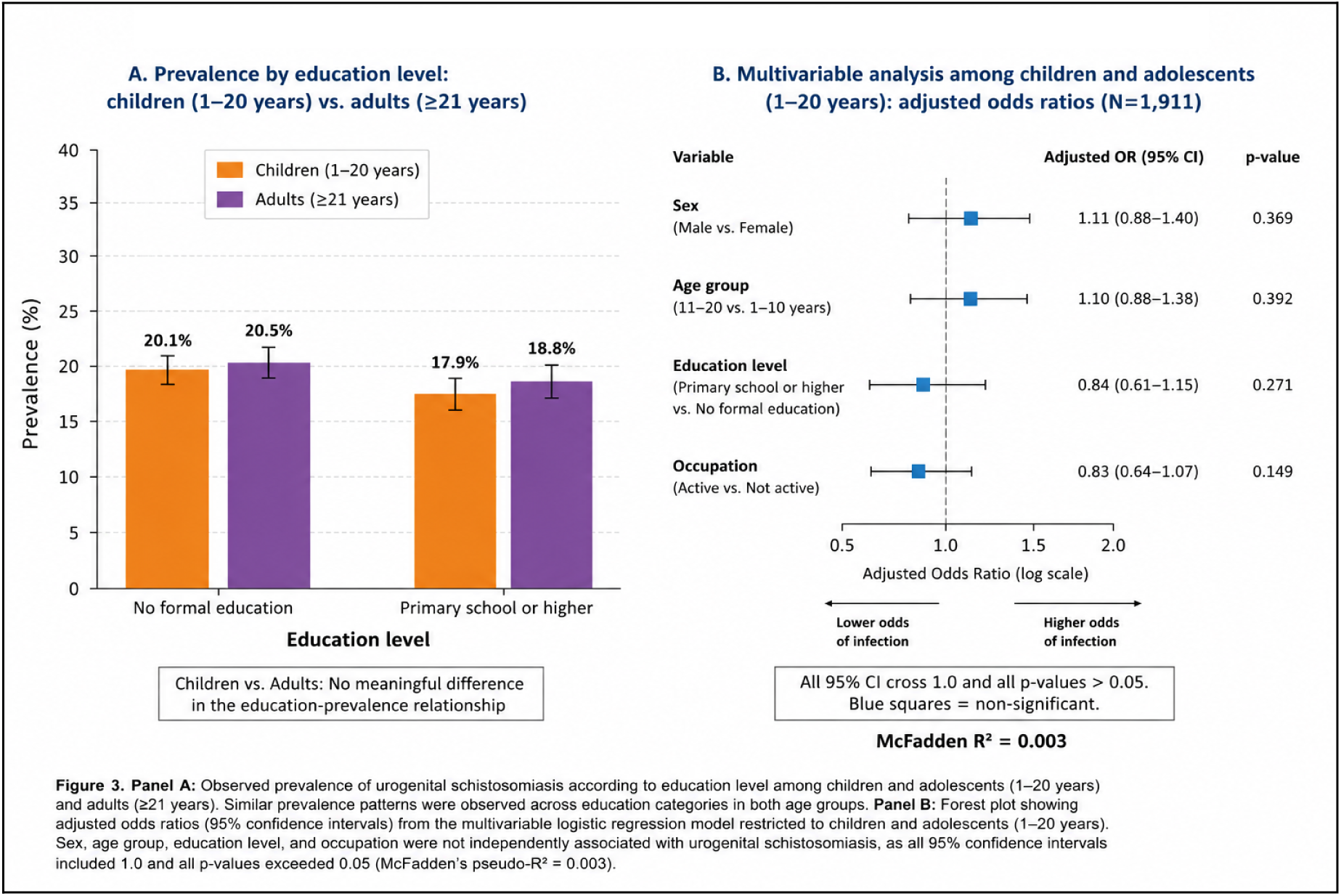
Determinants of urogenital schistosomiasis among children and adults. **Panel A:** Observed prevalence of urogenital schistosomiasis according to education level among children and adolescents (1–20 years) and adults (≥21 years). Similar prevalence patterns were observed across education categories in both age groups. **Panel B:** Forest plot showing adjusted odds ratios (95% confidence intervals) from the multivariable logistic regression model restricted to children and adolescents (N = 1,911). Sex, age group, education level, and occupation were not independently associated with urogenital schistosomiasis, as all 95% confidence intervals included 1.0 and all *p*-values exceeded 0.05 (McFadden’s pseudo-*R*² = 0.003)..

**Figure 4.**
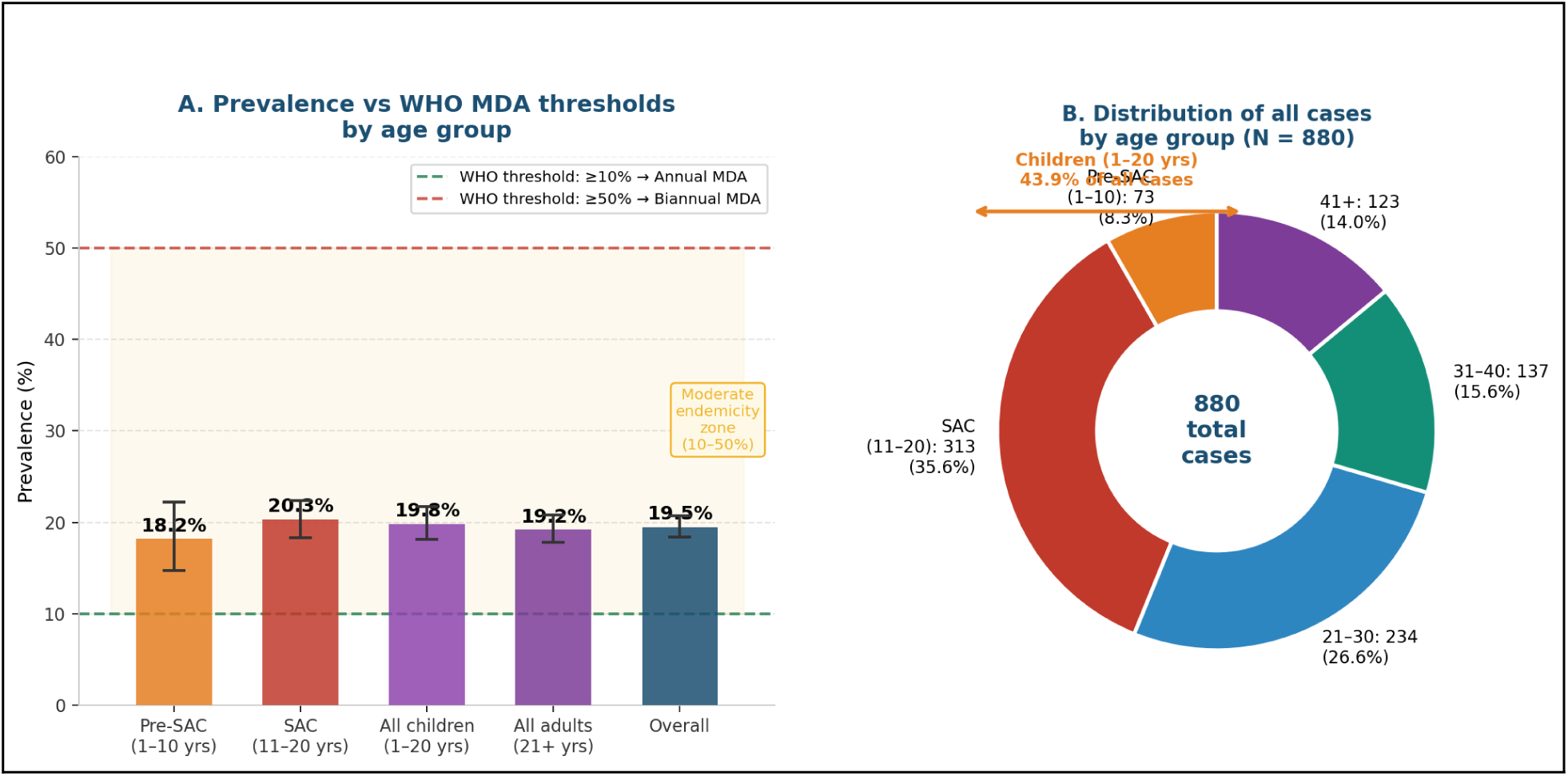
Prevalence relative to WHO MDA thresholds and case distribution by age group. Panel A: Parasitological prevalence by age group relative to WHO MDA thresholds. Both Pre-SAC (18.2%) and SAC (20.3%) exceed the 10% threshold for moderate endemicity, warranting annual MDA. The yellow shaded area represents the moderate endemicity zone (10%–50%). Panel B: Donut chart showing the distribution of all 880 confirmed cases by age group. Children and adolescents (1–20 years) account for 43.9% of total cases.

**Table 2.** WHO endemicity classification by age group and programmatic implications.

| Age group | Prevalence (%) | WHO endemicity class | WHO-recommended PC strategy |
| --- | --- | --- | --- |
| Pre-SAC (1–10 years) | 18.2 | Moderate (10–<50%) | Annual MDA — community-wide |
| SAC (11–20 years) | 20.3 | Moderate (10–<50%) | Annual MDA — community-wide |
| **All children (1–20 years)** | **19.8** | **Moderate (10–<50%)** | **Annual MDA — community-wide** |
| Adults (21+ years) | 19.2 | Moderate (10–<50%) | Annual MDA — community-wide |
| **Overall (all ages)** | **19.5** | **Moderate (10–<50%)** | **Annual MDA — community-wide** |

WHO thresholds: <10% low endemicity (no MDA indicated); 10%–<50% moderate endemicity (annual MDA); ≥50% high endemicity (biannual MDA). Source: WHO, 2022.

**Table 3.**
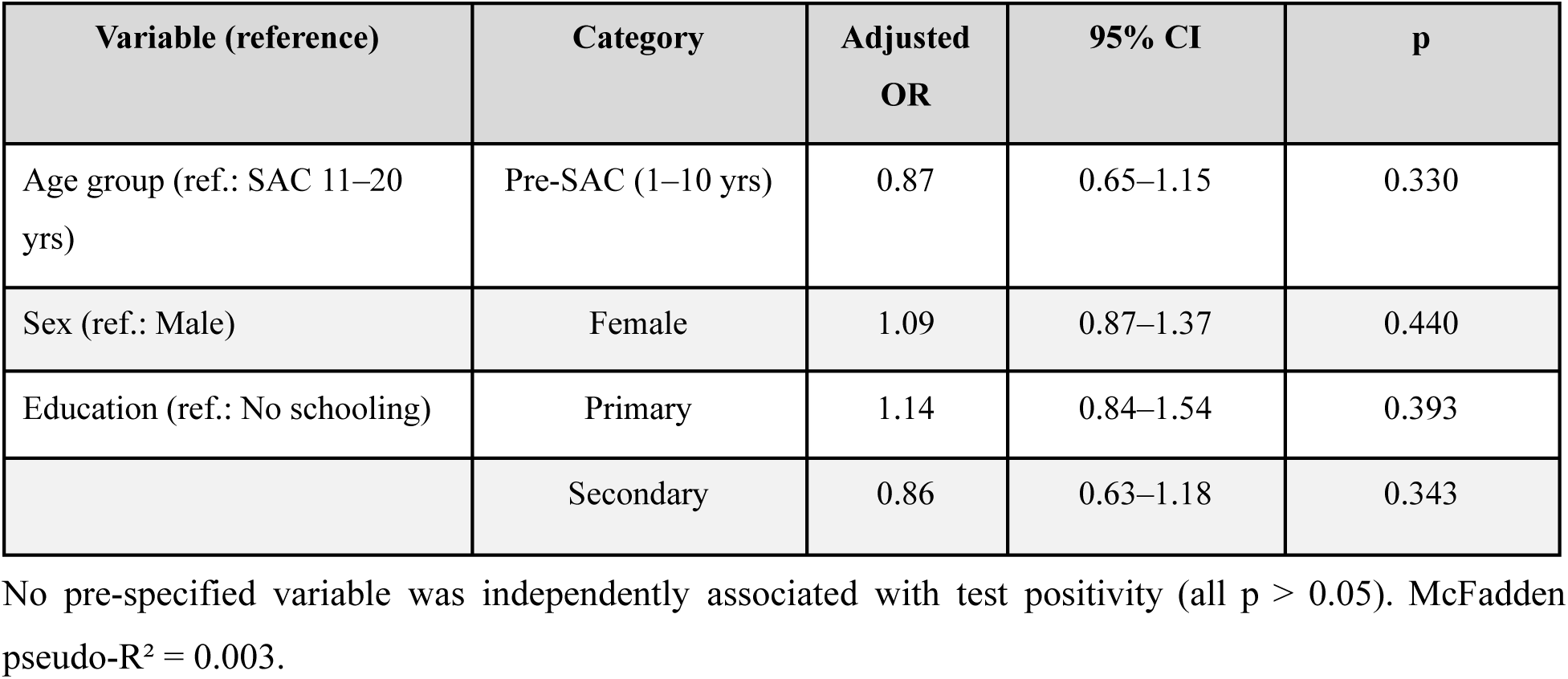
Multivariable logistic regression among children and adolescents (N = 1,911)

| Variable (reference) | Category | Adjusted OR | 95% CI | p |
| --- | --- | --- | --- | --- |
| Age group (ref.: SAC 11–20 yrs) | Pre-SAC (1–10 yrs) | 0.87 | 0.65–1.15 | 0.330 |
| Sex (ref.: Male) | Female | 1.09 | 0.87–1.37 | 0.440 |
| Education (ref.: No schooling) | Primary | 1.14 | 0.84–1.54 | 0.393 |
|  | Secondary | 0.86 | 0.63–1.18 | 0.343 |

No pre-specified variable was independently associated with test positivity (all $p > 0.05$ ). McFadden pseudo- $R^2 = 0.003$ .

## 5. DISCUSSION

### Persistent Community-Wide Transmission Across Age Groups

The principal finding of this study is that the observed prevalence of urogenital schistosomiasis during a 15-month community-based control programme remained remarkably similar across Pre-SAC children (18.2%), SAC adolescents (20.3%), and adults (19.2%). Furthermore, no demographic or clinical characteristic independently predicted infection among children after multivariable adjustment. Together, these findings suggest that transmission in the Ngouri district remains broadly distributed across the community rather than being concentrated within specific age groups.

Unlike many endemic settings in sub-Saharan Africa, where infection typically peaks during late childhood and adolescence because of age-specific water-contact behaviours, the epidemiological pattern observed here appears relatively homogeneous [1,11]. This likely reflects the agro-pastoral lifestyle of Lake Chad communities, where all household members are routinely exposed to the same contaminated water sources used for domestic activities, livestock watering, fishing and agriculture [6]. Consequently, transmission is maintained through shared environmental exposure rather than through behaviours restricted to school-aged children.

Importantly, these prevalence estimates should not be interpreted as the baseline epidemiological situation of the Ngouri district. Instead, they represent infection levels observed during routine programme implementation after 15 months of repeated community-based screening and treatment. The persistence of moderate endemicity despite these activities suggests that transmission remains active and that diagnosis and treatment alone may be insufficient to reduce infection below WHO programmatic thresholds.

### Pre-SAC Children Remain an Important Target Population

Pre-school and early school-age children (1–10 years) exhibited an observed prevalence of 18.2%, virtually identical to that of older children and adults. Although these estimates were obtained during programme implementation, they remain well above the WHO threshold requiring annual preventive chemotherapy [2]. This finding reinforces growing evidence that younger children constitute an important reservoir of infection in endemic communities [3,4].

In agro-pastoral settings such as the Lake Chad Basin, young children accompany their families during farming, water collection and livestock activities from an early age, resulting in frequent exposure to infested water long before school enrolment [6]. The comparable prevalence observed across age groups therefore supports the WHO recommendation to include Pre-SAC in preventive chemotherapy programmes rather than restricting treatment to school-attending children alone.

From a programmatic perspective, excluding Pre-SAC from MDA would leave a substantial number of infected children untreated. In the present analysis, 73 confirmed infections occurred among the 402 children aged 1–10 years, representing 8.3% of all detected cases. Although these figures reflect programme-period prevalence rather than baseline burden, they demonstrate that younger children continue to contribute meaningfully to the community reservoir of infection.

### Implications for Schistosomiasis Control Strategies

According to the 2022 WHO guideline, annual preventive chemotherapy is recommended when prevalence among school-aged children exceeds 10%, particularly when other high-risk groups are similarly affected [2]. Although the prevalence reported here reflects infection during programme implementation rather than baseline endemicity, both Pre-SAC and SAC remained within the WHO moderate-endemicity category after 15 months of community-based screening and treatment. These findings indicate that annual MDA remains justified in the Ngouri district.

More importantly, the persistence of moderate endemicity despite repeated diagnosis and treatment raises broader questions regarding current control strategies. Chemotherapy remains highly effective for reducing morbidity and parasite burden, but its impact on transmission may be limited where repeated exposure to contaminated water continues. Similar observations have been reported from several endemic African settings, where multiple rounds of MDA substantially reduced infection prevalence without interrupting transmission because environmental transmission sites remained unchanged.

These findings therefore argue for integrating preventive chemotherapy with complementary interventions, including improved water, sanitation and hygiene (WASH), health education, and more adaptive surveillance systems. Rather than relying exclusively on periodic prevalence surveys, dynamic hydro-epidemiological mapping capable of continuously identifying persistent transmission hotspots could improve targeting of interventions and optimise resource allocation. Integrating routine programme data with environmental information derived from remote sensing and geographic information systems represents a promising direction for future schistosomiasis control programmes.

### Comparison with Previous Studies

Direct comparison between the present findings and previous studies should be undertaken cautiously because the prevalence reported here was measured during programme implementation rather than before intervention. Consequently, these estimates are not directly comparable with baseline epidemiological surveys.

Nevertheless, the observed prevalence remains broadly consistent with previous work conducted in Chad. In Torrock (Mayo-Kebbi West), a community survey using the Dawa Mobile Health platform reported a prevalence of 24.9% among children aged 1–14 years [7]. The somewhat lower prevalence observed in the present analysis may partly reflect the cumulative effects of 15 months of community-based diagnosis and treatment. Unlike the Torrock study, however, we observed no significant difference in infection prevalence between boys and girls, suggesting that exposure patterns in Ngouri are more evenly distributed across sexes.

Likewise, the substantially higher prevalence reported in the Salamat region (55%) [12] most likely reflects a district with considerably higher baseline transmission before implementation of large-scale control activities. These comparisons highlight the importance of considering programme context when interpreting prevalence estimates across studies.

### Strengths and Limitations

This study has several important strengths. It includes one of the largest routinely collected schistosomiasis datasets currently available from Chad, encompassing 4,516 participants across all age groups, including 402 Pre-SAC children. Standardised parasitological diagnosis using WHO-recommended membrane filtration, systematic electronic data capture through the Dawa Mobile Health platform, and inclusion of both children and adults allowed comprehensive assessment of age-related infection patterns under real-world programme conditions.

The study also has important limitations. First, this was a secondary analysis of routinely collected programme data rather than a protocol-driven epidemiological survey. Consequently, the reported prevalence reflects infection levels observed during programme implementation and should not be interpreted as the underlying prevalence of the Ngouri district. Because screening and treatment occurred continuously throughout the 15-month implementation period, the observed prevalence is likely lower than the true pre-intervention prevalence.

Second, age was recorded in broad programme categories (1–10 and 11–20 years), preventing separate analysis of younger pre-school children (1–4 years) according to the strict WHO definition. Third, parasitological diagnosis relied on a single urine specimen, which may have underestimated infection because of day-to-day variation in egg excretion. Fourth, detailed information on individual water-contact behaviours and environmental exposure was unavailable, limiting exploration of the mechanisms underlying the observed homogeneous age distribution.

## CONCLUSION

This study provides a secondary analysis of routinely collected data from a 15-month community-based schistosomiasis control programme implemented in the Ngouri Health District, Chad. During programme implementation, the observed prevalence of urogenital schistosomiasis remained remarkably similar among Pre-SAC children (18.2%), school-age children and adolescents (20.3%), and adults (19.2%), with no sociodemographic or clinical characteristic independently associated with infection among children. These findings suggest that transmission is broadly distributed across the community rather than concentrated within specific demographic groups.

Importantly, the prevalence estimates reported here should not be interpreted as the baseline prevalence of the Ngouri district. Nevertheless, despite 15 months of community-based screening and treatment, the observed prevalence in both Pre-SAC and SAC remained above the WHO threshold requiring annual preventive chemotherapy, indicating persistent community transmission.

These findings support continued implementation of community-wide preventive chemotherapy, including explicit incorporation of Pre-SAC children into routine treatment programmes, together with sustained investments in water, sanitation and hygiene (WASH). However, they also suggest that chemotherapy alone may be insufficient to interrupt transmission in highly endemic agro-pastoral settings.

Future control strategies should therefore combine preventive chemotherapy with dynamic surveillance systems capable of continuously identifying residual transmission hotspots and guiding more targeted environmental and public health interventions. Integrating routine programme data with geospatial and environmental information may enable more adaptive and efficient schistosomiasis control, particularly in complex transmission settings such as the Lake Chad Basin.

Beyond their immediate programmatic implications, these findings contribute new evidence from Chad on age-related patterns of urogenital schistosomiasis under real-world programme conditions and reinforce the need for integrated control strategies that combine treatment, environmental interventions, and continuous epidemiological surveillance to accelerate progress towards schistosomiasis elimination.

## Data Availability

All data produced in the present study are available upon reasonable request to the authors

## Notes

### Competing Interest Statement

The authors have declared no competing interest.

### Author Declarations

The Dawa Mobile Health programme was approved by the Scientific Ethics Committee of Chad and the National Bioethics Committee of Chad before programme implementation.

### Funding

- Grand Challenges Canada: ST-POC-2312-62501

